# Peripheral regulatory CD8^+^CD28^−^KLRG1^+^ T cells as markers of disease and treatment response in rheumatoid arthritis

**DOI:** 10.1101/2020.09.02.20186635

**Authors:** Charlotte Thompson, Richard Beatson, Ruth Davies, Claire Greenhill, Simon A. Jones, Anwen S. Williams, Gareth W. Jones, Ernest HS Choy

## Abstract

**Objective:** CD3^+^CD8^+^CD28^−^ cells are increased in the periphery and tissues of rheumatoid arthritis (RA) patients. The aim of this study was to characterise CD3^+^CD8^+^CD28^−^ cells for the presence of cell surface receptors that regulate immune activation and function and to track their presence in a disease model of RA. Cell surface receptors expressed by CD3^+^CD8^+^CD28^−^ cells were then related to serological and clinical disease parameters to establish whether these cells are prognostic of a clinical response to conventional DMARDs.

**Method:** Using healthy donor peripheral blood mononuclear (PBMC) cell surface expression of > 50 candidate markers were tested using flow cytometry and compared against CD28 expression. The prevalence of cells expressing the most suitable candidate was investigated in the collagen induced arthritis (CIA) and the antigen-induced arthritis (AIA) models. Fifty RA patients were recruited from University Hospital of Wales (UHW) rheumatology outpatient clinic. Clinical and serological markers of inflammation were noted, and PBMC were analysed using flow cytometry +/− *in vitro* stimulation.

**Results:** CD3^+^CD8^+^CD28^−^ T cells express CD244, CD57, CX3CR1 and KLRG1. The strongest inverse correlate of CD28 expression was KLRG1. CD3^+^CD8^+^CD28^−^KLRG1^+^ cells were elevated in experimental models of RA. Notably, Il-10-deficiency was linked with exacerbated arthritis and an increase in the number of CD3^+^CD8^+^CD28^−^KLRG1^+^ cells, suggesting a regulatory role for Il-10 in their development or survival. In RA patients, CD3^+^CD8^+^CD28^−^KLRG1^+^ cells correlate with ACPA, RF and ESR, and produce more IL-10 than controls. Finally, these cells are higher in early arthritis patients that do not respond to treatment with synthetic DMARDs at six months.

**Conclusion:** KLRG1 is a marker for regulatory CD3^+^CD8^+^CD28^−^ cells. The presence of CD3^+^CD8^+^CD28-KLRG1^+^ cells increases with certain measures of disease, and is indicative of poor treatment response to DMARDs in early arthritis.

**Key Messages:**

1. KLRG1 is a marker of CD28 negativity on CD8 T cells
2. CD3^+^CD8^+^CD28^−^KLRG1^+^ cells are increased in CIA mice and correlate with disease severity.
3. CD3^+^CD8^+^CD28^−^KLRG1^+^ cells positively correlate with ESR / ACPA and RF in patients.
4. CD3^+^CD8^+^CD28^−^KLRG1^+^ cells are increased in Il-10-deficient mice with inflammatory arthritis.
5. CD3^+^CD8^+^CD28^−^KLRG1^+^ cells produce more Il-10 than CD3^+^CD8^+^CD28^+^KLRG1^−^cells in RA patients.
6. CD3^+^CD8^+^CD28^−^KLRG1^+^ cells are higher in patients who do not respond to treatment with synthetic DMARDs after six months.

## Introduction

CD3^+^CD8^+^ T lymphocytes are raised in the periphery of RA patients in comparison to healthy controls^1^. These cells are also found in abundance in the synovial fluid of patients with RA ^2^. CD3^+^CD8^+^ T cells can be divided into the greater pro-inflammatory and lesser regulatory cell populations^1,3^. Increased pro-inflammatory cytokine production by CD3^+^CD8^+^ effector T cells in peripheral blood of patients with RA, normalises in remission^1^.

CD28 is a pivotal cell surface marker on CD3^+^CD8^+^ T cells. When CD28 is expressed, the T cell plays a predominantly pro-inflammatory role. A loss of CD28 occurs during antigen-driven differentiation toward an effector memory (EM) or terminal effector (TEMRA) phenotype. Intriguingly, when CD28 is lost, T cells have been reported to have more of an immunosuppressive role in cancer, transplantation and systemic lupus erythematosus (SLE) ^3^,^4^,^5^. CD3^+^CD8^+^CD28^−^ cells are known to be raised in RA ^6,7^, however, they have lost the ability to immunosuppress autologous lymphocyte proliferation^7^. Tumor necrosis factor inhibitors (TNFi) can partially restore their immunosuppression and transwell experiments suggested both soluble factors and direct cell surface contact contribute to their function^7^.

In order to better understand the role of CD8+CD28- T cells in RA we wished to find a positive marker that associated with function, disease and prognosis.

## Methods

### Preparation of human peripheral blood mononuclear cells (PBMCs), murine spleen mononuclear cells and lymph node cells

Fresh healthy peripheral blood was obtained from healthy volunteers, via the NHS blood transfusion service, between 2011 and 2012. RA patient blood was obtained between 2014 and 2017. PBMCs were separated from blood using Ficoll-paque and PBMCs were used immediately. Murine spleens were squeezed through nylon mesh. Mononuclear cells were separated on Ficoll-paque (GE Healthcare) before being stained immediately. Lymph node cells were isolated in the same manner, omitting density gradient separation.

### Flow Cytometry

All cells were assessed for viability using a fixable viability dye (ebioscience eFluor506). Flow cytometric analysis of human and murine cells was performed using > 50 antibodies and appropriate directly conjugated mouse, rat and hamster isotype controls. The clones relevant to the majority of work are as follows: mouse anti-CD3-FITC (Miltenyi; REA641) anti-CD8a-PerCP (Miltenyi; 53–6.7) anti-CD28-PE (Miltenyi; 37.51) anti-KLRG1-APC (Miltenyi; 2F1). Human anti-CD3 APC-Cy7 (biolegend; HIT3a) anti-CD8-PE-Vio770 (Miltenyi; REA734) anti-CD28-eFluor450 (ebioscience; CD28.2) anti-KLRG1-AF488 (a generous gift from Professor Hanspeter Pircher, University of Freiburg) anti-CD57-Vioblue (Miltenyi; TB03) anti-CD244-FITC (biolegend; C1.7) anti-CX3CR1-PE (ebioscience; 2A9–1), and anti-IL-10-FITC (ebioscience; BT-10). For intracellular cytokine staining, cells were cultured with PMA (50 ng/ml), ionomycin (500 ng/ml), and monensin (3 µM; all from Sigma) for 4 h at 37C. Cells were stained for cell surface markers, then fixed and permeabilized in Cytofix/Cytoperm (BD) before intracellular detection of cytokines. Cells were acquired using a CyAn ADP (Beckman Coulter) flow cytometer and analysed using Summit software (software version 4.3; Beckman Coulter). Percentages and mean fluorescence intensity (MFI) were calculated against appropriate isotype controls.

### Isolation of human and mouse CD8+CD28-KLRG1+ cells

CD8 cells from human PBMCs from fresh blood were isolated by negative selection using the CD8^+^ T cell isolation kit (Miltenyi Biotech) as per manufacturer’s instructions. CD8^+^CD28^−^ cells were then isolated by negative selection using CD28 microbeads (Miltenyi Biotech) as per manufacturer’s instructions. CD8^+^KLRG1^+^ cells from murine splenocytes were isolated by positive selection using first, CD8a microbeads (Miltenyi Biotech) and, using a second isolation, KLRG1 microbeads (Miltenyi Biotech). Cells were used immediately.

### Proliferation assays

96 well plates were coated with human (OKT-3; Biolegend) or mouse anti-CD3 (17A2; Miltenyi Biotech) depending of the planned assay on day 0. On day 1 PBMCs or isolated cell subsets were stained with a cell proliferation dye (eFluor450; ebiosciences) as per manufacturer’s instructions. For proliferation assays, labelled isolated cell subsets were plated at 0.5×10^6^/ml (200µl) on anti-CD3, or with CD3/CD28 microbeads (ThermoFisher), or with 5µg/ml PHA (Sigma). Cells were incubated at 37C for 3 days before being analysed by flow cytometry. For inhibition assays, labelled PBMCs were plated at 1×10^6^/ml (100µl) on anti-CD3 before different indicated concentrations of CD8^+^CD28^−^ cells (human) or CD8^+^KLRG1^+^ cells (mouse) were added. Cells were cultured for 48h before being analysed using flow cytometry.

### Collagen-Induced Arthritis

Experiments were undertaken in 8-week-old male DBA-1 mice. Mice were sourced from Charles River, UK. Procedures were performed in accordance with home office approved project license 30/2928 and personal licence I5/6CC73C0. In brief, mice were immunized on two occasions, 21 days apart, with identical 100 µL intradermal injections of an emulsion containing 1 mg/mL type II chicken sternal collagen (Sigma) and 2.5 mg/mL Freund’s complete adjuvant. Temgesic (0.4 mg/mL) was administered *ad libitum* via the drinking water on day 20 before arthritis onset and was continued until the end of each experiment. Arthritis progression was monitored and scored manually 0 = normal, 1 = redness/swelling or both in one joint, 2 = redness/swelling or both in more than one joint, 3 = redness/swelling or both in the entire paw, and 4 = deformity/ankylosis or both.

### Antigen-Induced Arthritis

Inbred C57BL/6 mouse strains from Charles River were bred and maintained in-house under high barrier and pathogen-free conditions. Experiments were performed on 8–12 week old male mice in accordance with UK Home Office Project License PPL-30/2361 and 30/2928. Mice were immunized (subcutaneous (s.c.)) with an emulsion containing 1 mg/ml methylated bovine serum albumin (mBSA) in phosphate-buffered saline (PBS) and Freud’s complete adjuvant (CFA) (Sigma). Concurrently, mice were injected (intraperitoneal (i.p.)) with 200ng of heat-inactivated *Bordetella pertussis* toxin adjuvant (Sigma). The immune response was boosted one week later with a second injection (s.c.) of mBSA emulsified in CFA. Arthritis was induced two weeks later with an intra-articular (i.a.) injection of 10μl of mBSA (10 mg/ml) into the right knee joint. Arthritis progression was monitored using a micrometer to measure changes in knee joint swelling.

### Patients

25 patients were recruited from Cardiff University Hospital of Wales (UHW) rheumatology outpatient clinic to the Early Arthritis study arm, 25 to the Established Arthritis arm and 25 Healthy Controls. The study was approved by the South East Wales Research Ethics Committee, Panel B in 2011 (REC reference: 11/WA/0326). Cardiff University was responsible for the governance of the study with reference number 11/CMC/5299. The Cardiff and Vale University Health Board Research & Development Office approved the proposal in 2013. Inclusion criteria included the following: age ≥18 years; meets the criteria of 2010 RA diagnosis; synovitis in at least one joint; duration of persistent symptoms in the Early RA group of 4 weeks to 6 months, > 2 years since diagnosis for the Established RA group^8,9,10^. Exclusion criteria include the following: definite other autoimmune rheumatic disease; heart disease classified as New York Heart Foundation Functional class IV (ACR classification); treatment with intravenous gamma globulin, plasmapheresis or Prosorba^TM^ column within the last 6 months; current other inflammatory joint disease.

### Clinical Assessments

During baseline clinical assessments, the following was data was collected: age, gender, RA disease duration, DAS28 score, RF, ACPA, ESR and CRP^11^. Demographic data and clinical characteristics are shown in Table 1.

**Table 1.** Demographics of health controls, early and established RA patients recruited. Percentages are shown with number of subjects in brackets. Mean average is presented. Age and DAS28 are shown with standard deviation.

|  | Healthy Controls<br>(n=25) | Early RA<br>(n=25) | Established RA<br>(n=25) |
| --- | --- | --- | --- |
| Age | 41±12 | 56±12 | 62±12 |
| Disease duration | N/A | 4 Months | 10 Years |
| RF Positive (%) | N/A | 68 (17) | 56 (14) |
| ACPA positive (%) | N/A | 79 (19) | 63 (15) |
| DAS28 | N/A | 4.07±1.3 | 5.4±1.9 |
| Female (%) | 68 (17) | 64 (16) | 68 (17) |

### Statistics

Statistical analysis was performed using GraphPad Prism v5 and v8 software. Significant differences were determined using the nonparametric Mann Witney U Test or the parametric Student’s t-test, both paired and unpaired, with Welch’s correction where appropriate. The comparison of the difference between three or more unmatched groups was established using the Kruskal-Wallis test was used. p< 0.05 was considered significant. The Spearman rank correlation was used to analyse statistical associations and presented with a line of linear regression.

## Results

### KLRG1 is a positive marker for CD8^+^CD28^−^ regulatory T cells

To define a positive marker of CD28 negativity, peripheral blood mononuclear cells (PBMCs) were isolated from healthy donors and stained for CD3, CD8, CD28 and a > 50 potential CD28 negative correlates using flow cytometry (data not shown). Four markers were of sufficient intensity and fold change to take forward: CD244, CD57, CX3CR1 and KLRG1. Linear regression analysis was performed on each marker’s expression against CD28 expression (Figure 1A-D). As KLRG1 expression had the strongest inverse relationship with CD28 expression, it was chosen as the positive marker.

We next assessed the function of CD8^+^CD28^−^KLRG1^+^ cells finding that they proliferated well in response to PHA and anti-CD3 stimulation (Figure 1E). We were able to demonstrate that CD8^+^CD28^−^KLRG1^+^ cells from both healthy humans and mice were able to inhibit autologous PBMC proliferation (Figure 1F,G). These assays demonstrated that in healthy humans and mice, CD8^+^CD28^−^KLRG1^+^ cells defined a regulatory phenotype.

### CD3^+^CD8^+^CD28^−^KLRG1^+^cells are increased in the spleens and inguinal lymph nodes of CIA and AIA mice, correlating with disease severity

To assess if CD3^+^CD8^+^CD28^−^KLRG1^+^ may be an appropriate marker of disease in RA patients, we used the well-characterised CIA and AIA models. CD3^+^CD8^+^CD28^−^KLRG1^+^ cells were increased in the spleens in both models compared to healthy littermate controls (Figure 1H), and intriguingly increased further in an *Il10^−/-^* model using the AIA model in line with a previously reported increase in disease severity (Figure 1I). The CIA model was then assessed for correlations of disease score and % CD8^+^CD28^−^KLRG1^+^ cells the spleens and inguinal lymph nodes, finding significant correlations in both instances (Figure 1J,K). Therefore we considered splenic regulatory CD8^+^CD28^−^KLRG1^+^ cells to be a readout of disease severity in these models.

**Figure 1.**
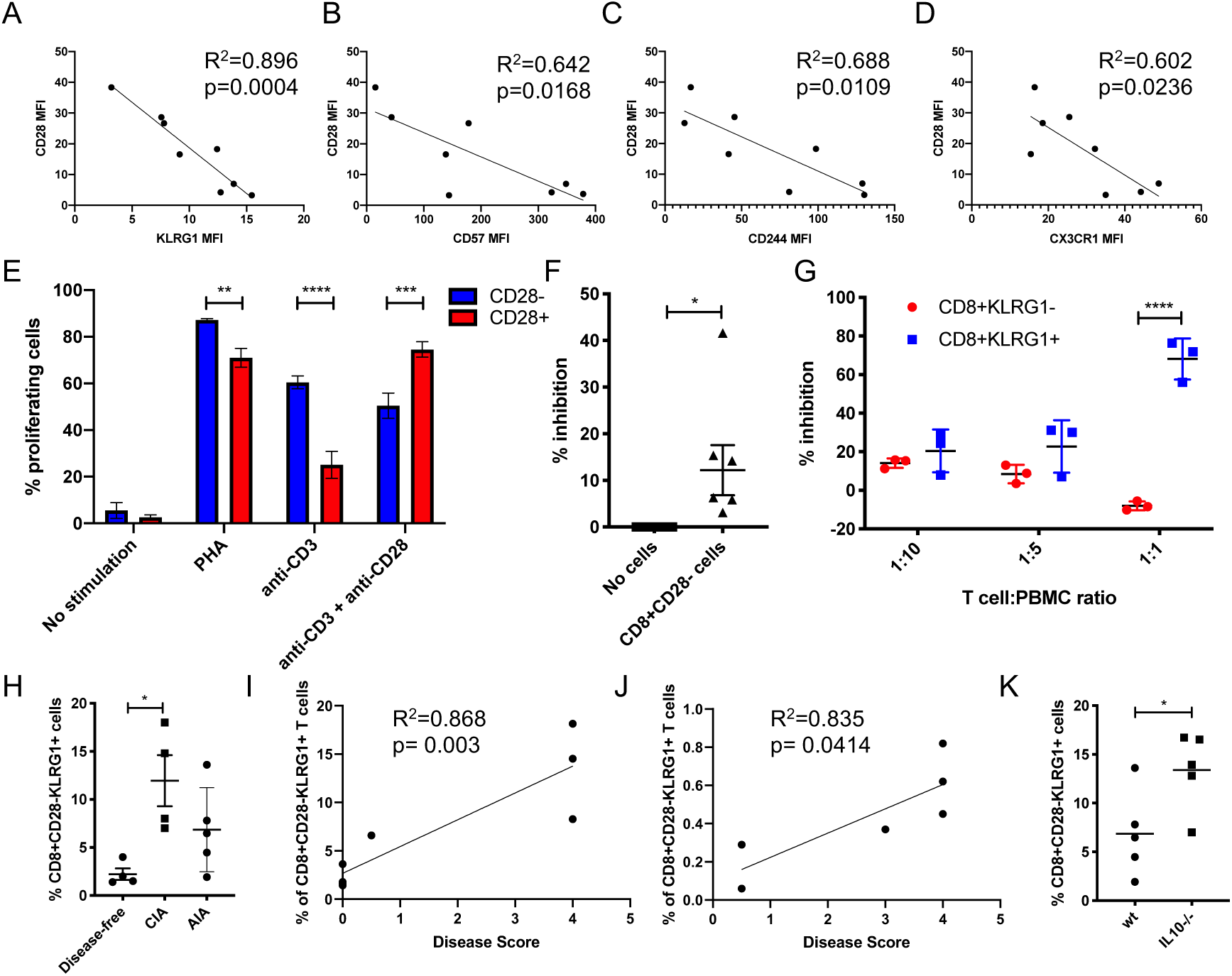
KLRG1 is a marker of CD28 negativity on CD8 T cells, with CD8+CD28-KLRG1+ T cells correlating with disease severity in murine models. A-D) Linear regression analysis between CD28 MFI and indicated MFI (A; KLRG1. B; CD57. C; CD244. D; CX3CR1) of CD8 T cells (n = 8). Spearman’s coefficient and p value shown. E) % of proliferating indicated cells after stimulation with indicated factors for 72h (n = 3). Paired t test. F) % inhibition of CD3 stimulated PBMC proliferation in the presence or absence of autologous CD8^+^CD28^−^ T cells (1:1 ratio; n = 7). Paired t test. G) % proliferating CD3 stimulated PBMC when co-cultured with indicated ratios with autologous indicated cells (n = 3). Paired t test. H) Grouped analysis of % CD8^+^CD28-KLRG1^+^ cells within total splenocytes, in AIA mice, CIA mice and healthy litter-matched (with CIA) controls (n = 4–5). Unpaired t-test. I-J) Linear regression analysis between disease score and percentage of CD3^+^CD28-KLRG1^+^ cells in the spleen (I) and inguinal lymph nodes (J) in littermate control and CIA mice (n = 4). Spearman’s coefficient and p value shown. K) CD8^+^CD28^−^KLRG1^+^ cells in the spleens of wt or Il10^−/-^ AIA mice (n = 5). Unpaired t-test. Mean and SEM shown. *p< 0.05 **p< 0.01, ***p< 0.001, ****p< 0.0001.

### Peripheral CD3^+^CD8^+^CD28^−^KLRG1^+^ T cells positively correlate with measures of disease, produce IL-10 and correlate with DMARD refractory disease in early RA

Given our murine data we wished to assess whether the presence of CD8^+^CD28^−^KLRG1^+^ cells correlated with disease severity in RA patients. Demographics and clinical groupings of the RA patient cohorts can be found in Table 1. First, we assessed if there was an increase in CD3^+^CD8^+^CD28^−^KLRG1^+^ cells in early or established disease. Percentage of CD3^+^CD8^+^CD28^−^KLRG1^+^ in healthy controls was numerically higher in RA patients 30%+22% compared with 25%+16% in healthly controls (p = 0.052) (Figure 2A). Early (33%+19%) and established RA (37%+25%) patients have similar percentage of CD3^+^ CD8^+^CD28^−^KLRG1^+^ cells. Next, we investigated clinical measures of disease activity. There was a significant correlation between the percentage of peripheral CD3^+^CD8^+^CD28-KLRG1^+^ T cells and ESR in the pooled RA and early RA patient groups (Figure 2B,C). The percentage of CD3^+^CD8^+^CD28^−^KLRG1^+^ cells in the peripheral blood were found to be higher in RF positive (Figure 2D) and ACPA positive (Figure 2E) established RA patients. However there was no statistically significant correlation between peripheral CD3^+^CD8^+^CD28^−^KLRG1^+^ T cells and CRP, age, DAS28 or disease duration (data not shown). This collective data suggested that these cells could not be used as a single marker of overall disease severity.

**Figure 2.**
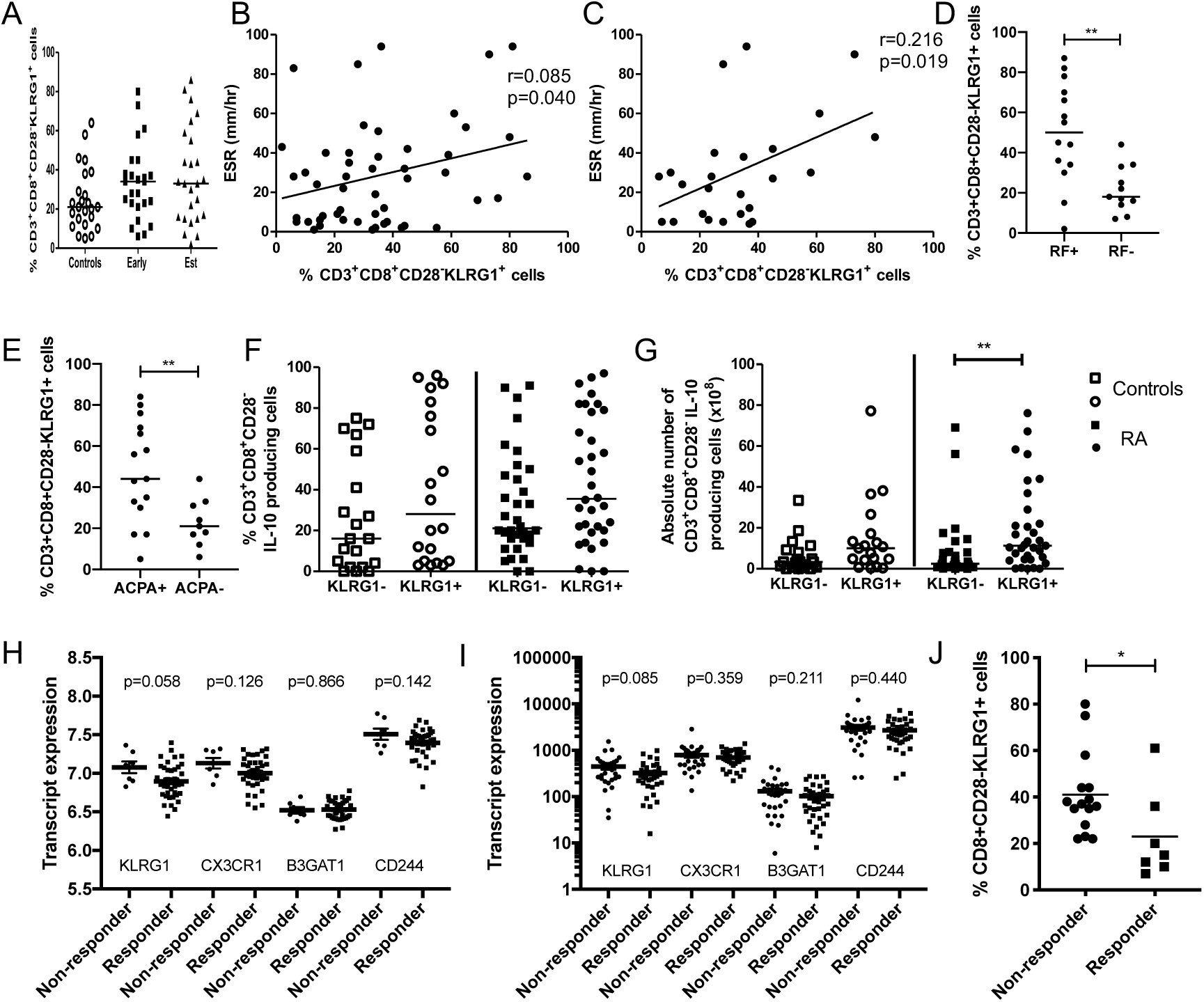
Percentage of CD3^+^CD8^+^CD28^−^KLRG1^+^ cells correlate with disease parameters, produce IL-10 and are indicative of treatment response to DMARDs. A) %CD3^+^CD8^+^CD28^−^KLRG1^+^ cells in PBMCs of healthy donors, early or established RA patients (n = 7, 25, 25 respectively). B-C) Linear regression analysis of ESR and percentage of CD3^+^CD8^+^CD28^−^KLRG1^+^ cells in PBMCs of pooled (B) and early (C) RA patients. Pooled RA (n = 50), early RA (n = 25). Spearman’s coefficient and p value shown. D-E) Percentage of CD3^+^CD8^+^CD28^−^KLRG1^+^ cells in PBMCs of RA patients with RF (D) or ACPA (E) positive or negative disease. Bars represent median, Mann-Witney U Test. F-G) Percentage (F) and absolute number (G) of CD3^+^CD8^+^CD28^−^KLRG1^+^IL-10^+^ cells in RA and control PBMCs after *in vitro* stimulation. Bars represent median. Kruskal-Wallis ANOVA test. Controls (n = 19), Pooled RA (n = 33). H-I) Expression levels of indicated transcripts in whole blood from RA patients who responded to iTNF biologics (responder) and those who did not respond (non-responder). Array (H) and RNAseq (I) data taken from Julia et al. (responders; 37. Non-responders; 7) and Farutin et al. (responders; 36. Non-responders; 27) respectively. Mean and SEM shown. Unpaired t test. J) Percentage of CD3^+^CD8^+^CD28^−^KLRG1^+^ cells in PBMCs of DMARD responders or non-responders in early arthritis at 6 months. Bars represent median. Mann-Witney U Test. Three patients were excluded because they did not have a recorded DAS28 at six months. Early RA (n = 22, responders = 7, non-responders = 15). *p< 0.05 **p< 0.01.

KLRG1+ cells have previously been reported to be associated with IL-10 in RA, and CD8^+^CD28^−^ T cells have been reported to secrete IL-10 to regulate T cell proliferation. To explore this relationship we assessed for IL-10 producing cells in RA patients, finding that the percentage and absolute number of peripheral CD3^+^CD8^+^CD28^−^KLRG1^+^ cells producing IL-10 in pooled RA was higher than in CD3^+^CD8^+^CD28^−^KLRG1^−^ cells (Figure 2F,G). This along with our AIA mouse data suggests that IL-10 may play a role in the function or production/maintenance of these cells.

We next questioned if CD3+CD8+CD28-KLRG1+ cells may be altered in patients who respond to therapy. We assessed KLRG1, CX3CR1, B3GAT1 (the glycotransferase that is most important in the production of the glycan, CD57) and CD244 expression levels in patients who responded, and did not respond, to TNFi therapy using publicly available whole blood datasets. We found that both datasets showed a decrease in all of the CD28 negative correlates in responders, however only KLRG1 was close to significance. In our own cohort, when assessing disease response to synthetic DMARDs, the percentage of peripheral CD3+CD8+CD28-KLRG1+ T Cells is higher in early RA non-responders than responders (Figure 2). Response to treatment was determined as a DAS28 reduction of > 1.2.

## Discussion

CD3^+^CD8^+^CD28^−^ cells classically contain both the effector memory (EM) and CD45RA+ terminal effector memory (TEMRA) populations. This mixed population is associated with disease in RA however robust positive markers that provide insight into the functions of these mixed phenotypes are lacking. By adopting a literature-led proteomic approach and assessing > 50 potential cell surface markers this study identified 4 markers of CD28 negativity in healthy donors: CD244, CX3CR1, CD57 and KLRG1. It is important to emphasise that these markers are not synonymous with CD28 negativity, however the strength of their correlations reveals that they represent significant populations within this group.

CD244 is most commonly considered a costimulatory receptor expressed on NK cells and activated T cells. It binds to CD48 on T cells and inflamed tissue leading to enhanced cytokine release and cytotoxic effects. CX3CR1, or fractalkine receptor, is a chemokine and adherence receptor for CX3CL1, expressed on activated lymphoid cells and monocytes. CX3CL1 is expressed in the brain and inflammatory sites. CD57 is a glycan (key glycosyltransferase; B3GAT1) associated with immunological aging, senescence and increased responsiveness. It is most commonly found on NK and T cells and has been reported to be associated with CD28 negatively in many publications. KLRG1 is found on NK and T cells with its expression being associated with high level of differentiation, immunosenescence and inhibitory function. Its ligands are E and N-cadherin.

These markers collectively are considered to be indicators of antigen experience, ‘NK drift’ and exhaustion, indeed CD8^+^CD28^−^ and TEMRA T cells have been reported to be senescent and unresponsive. Intriguingly three of these four markers bind ligands expressed on inflamed tissue, and this coupled with their proliferative potential suggests that rather than being senescent, these cells can expand through TCR stimulation alone in inflammatory environments. This may explain their increased prevalence in diseases such as RA; a result of active expansion of a specific phenotype in a specific disease setting, rather than exhaustion, senesence and lack of clearance.

To assess the presence of CD3^+^CD8^+^CD28^−^KLRG1^+^ cells in disease we firstly assessed for their presence in appropriate animal models finding an increase of these cells in both the spleen and inguinal lymph nodes that correlated with disease severity. However in the human disease setting using peripheral blood, we found a mixed picture. RF, ACPA and ESR, which are associated with a poor prognosis in RA^12^, correlated with the percentage of CD3^+^CD8^+^CD28^−^KLRG1^+^ cells. We speculate that RF and ACPA contribute to the repeated immune stimulation (these autoantibodies can directly stimulate macrophages and activate complement^12,13,14^) resulting in the development of CD3^+^CD8^+^CD28^−^KLRG1^+^ cells. The increase in inflammation would, in turn, increase the ESR, which also correlates with CD3^+^CD8^+^CD28^−^KLRG1^+^ cells.

KLRG1 was chosen as the marker that was most suitable for our study as its cell surface expression was high, it was significantly upregulated in the CD28- population and its expression was the most significant negative correlate with CD28 expression. Additionally, other groups have observed similar results^15^, and E-cadherin, both secreted and membrane-bound, has been reported to be raised in RA^10^. In line with other reports assessing CD8^+^CD28^−^ cell function, we found that CD8^+^CD28^−^KLRG1^+^ cells can inhibit autologous PBMC proliferation in human and murine systems.

The literature suggests that IL-10 may be involved in the immunoregulatory function and maintenance of these cells, including the interaction of KLRG1 with its ligand, and so we first assessed if the presence of these cells was altered in *Il10*^−/-^ mice using the AIA model. We found these cells were significantly increased in the spleen of *Il10*^−/-^ animals suggesting that these cells were not induced in response to Il-10, however this assay could not inform us whether CD3^+^CD8^+^CD28^−^KLRG1^+^ cells were a readout of disease enhancement (*Il10^−/-^* enhanced pathology^16^) and greater antigen experience, or whether, as hypothesised producers of IL-10 and, given the known sensitivity of CD8 T cells to IL-10, these cells were expanding owing to the absence of inhibitory autocrine signalling. To help answer this question, RA PBMCs were stimulated *ex vivo* and stained for IL-10. The increased production of IL-10 from this subset of cells suggested that they may control inflammation through this cytokine. It additionally strengthens the possibility that the IL-10 produced by these cells inhibits their proliferation in an autocrine fashion, and as such they dynamically self-regulate in line with disease. Other cytokines including IFN-g, IL-6, IL-4 and IL-17A were investigated; differences were not significant.

Finally, as producers of IL-10 and potential ‘read-outs’ and regulators of inflammation in RA, we assessed whether the presence of CD3^+^CD8^+^CD28^−^KLRG1^+^ T cells was associated with treatment response. We first assessed publicly available datasets, finding that even in whole blood, KLRG1 was higher, almost to a significant level in patients that did not respond to biological therapy. In our cohort we similarly found a significantly higher presence of CD3^+^CD8^+^CD28^−^KLRG1^+^ cells in patients that have not responded to synthetic DMARDs after six months. The increase, or lack of decrease, could indicate a greater inflammatory burden beyond these cells’ resolving capacities, or a form of inflammation that is refractory to IL-10.

The collective data suggests that KLRG1 can be used as a marker to identify a subset of resolving CD8 T cells that are increased in patients with RA. Furthermore, a decrease in these cells is associated with therapeutic response. CD8^+^CD28^−^KLRG1^+^ cells appear to represent a population of pro-resolving CD8 cells that fail to counteract a high inflammatory state in RA. Learning how to enhance the activities of these may have clinical benefit. KLRG1, as a marker of refractory disease, could therefore warrant a higher degree of intense treatment with DMARDs or, potentially, biologics. Further studies assessing the biology of CD8^+^CD28^−^KLRG1^+^ T cells would aid in their clinical utilisation, initially as a biomarker of treatment response, but potentially as a population that could be enhanced for therapeutic gain.

## Data Availability

All data (for the patient data, subject to ethical approval) is available upon request.

## Acknowledgments

CREATE Centre is funded by Versus Arthritis and Health and Care Research Wales.

## References

1 Carvalheiro H, Duarte C, Silva-Cardoso S, da Silva JA, Souto-Carneiro MM. CD8+ T cell profiles in patients with rheumatoid arthritis and their relationship to disease activity. Arthritis Rheumatol. 67, 363–371 (2015).

2 Cho BA Sim JH, Park JA, Kim HW, Yoo WH, Lee SH, Lee DS, Kang JS, Hwang YI, Lee WJ, Kang I, Lee EB, Kim HR. Characterization of effector memory CD8+ T cells in the synovial fluid of rheumatoid arthritis. J. Clin. Immunol. 32, 709–720 (2012).

3 Filaci G, Fenoglio D, Fravega M, Ansaldo G, Borgonovo G, Traverso P et al. CD8+ CD28- T regulatory lymphocytes inhibiting T cell proliferative and cytotoxic functions infiltrate human cancers. J Immunol. 2007 10 1; 179(7):4323–34.

4 Colovai A, Mirza M, Vlad G, Wang Su, Ho E, Cortesini R, Suciu-Foca N. Regulatory CD8+CD28− T cells in heart transplant recipients. Hum Immunol. 2003 Jan;64(1):31–7.

5 Tulunay A, Yavuz S, Direskeneli H, Eksioglu-Demiralp E. CD8+CD28−, suppressive T cells in systemic lupus erythematosus. Lupus. 2008 Jul; 17(7):630–7.

6 Sfikakis PP, Zografou A, Viglis V, Iniotaki-Theodoraki A, Piskontaki I, Tsokos GC, Sfikakis P, Choremi-Papadopoulou H. CD28 expression on T cell subsets in vivo and CD28-mediated T cell response in vitro in patients with rheumatoid arthritis. Arthritis Rheum. 1995 May;38(5):649–54.

7 Ceeraz S, Hall C, Choy EH, Spencer J, Corrigall VM. Defective CD8+CD28+ regulatory T cell suppressor function in rheumatoid arthritis is restored by tumour necrosis factor inhibitor therapy. Clin Exp Immunol. 2013 Oct;174(1):18–26.

8 Aletaha D, Neogi T, Silman AJ, Funovits J, Felson DT, Bingham CO, Birnbaum NS, Burmester GR, Bykerk VP, Cohen MD, Combe B, Costenbader KH, Dougados M, Emery P, Ferraccioli G, Hazes JM, Hobbs K, Huizinga TW, Kavanaugh A, Kay J, Kvien TK, Laing T, Mease P, Ménard HA, Moreland LW, Naden RL, Pincus T, Smolen JS, Stanislawska-Biernat E, Symmons D, Tak PP, Upchurch KS, Vencovský J, Wolfe F, Hawker G. 2010. Rheumatoid arthritis classification criteria: an American College of Rheumatology/European League Against Rheumatism collaborative initiative. Ann Rheum Dis. 69:1580–8.

9 TaylorJC, Bongartz T, Massey J, Mifsud B, Spiliopoulou A, Scott IC, Wang J, Morgan M, Plant D, Colombo M, Orchard P, Twigg S, McInnes IB, Porter D, Freeston JE, Nam JL, Cordell HJ, Isaacs JD, Strathdee JL, Arnett D, de Hair MJH, Tak PP, Aslibekyan S, van Vollenhoven RF, Padyukov L, Bridges SL, Pitzalis C, Cope AP, Verstappen SMM, Emery P, Barnes MR, Agakov F, McKeigue P, Mushiroda T, Kubo M, Weinshilboum R, Barton A, Morgan AW, Barrett JH; Matura; and Pamera; Consortia. Genome-wide association study of response to methotrexate in early rheumatoid arthritis patients. Pharmacogenomics J. 2018 May 25. doi:10.1038/s41397-018-0025-5.

10 Emery P, Breedveld FC, Hall S, Durez P, Chang DJ, Robertson D, Singh A, Pedersen RD, Koenig AS, Freundlich B. Comparison of methotrexate monotherapy with a combination of methotrexate and etanercept in active, early, moderate to severe rheumatoid arthritis (COMET): a randomised, double-blind, parallel treatment trial. Lancet. 2008 Aug 2;372(9636):375–82. doi: 10.1016/S0140-6736(08)61000-4. Epub 2008 Jul 16.

11 Svensson B, Schaufelberger C, Teleman A, Theander J. Remission and response to early treatment of RA assessed by the disease activity score. Rheumatology 2000;39:1031–6.

12 Trouw LA, Haisma EM, Levarht EW, van der Woude D, Ioan-Facsinay A, Daha MR, et al. Anti-cyclic citrullinated peptide antibodies from rheumatoid arthritis patients activate complement via both the classical and alternative pathways. Arthritis Rheum. (2009) 60:1923–31. 10.1002/art.24622

13 Lu MC, Lai NS, Yu HC, Huang HB, Hsieh SC, Yu CL. Anti-citrullinated protein antibodies bind surface-expressed citrullinated Grp78 on monocyte/macrophages and stimulate tumor necrosis factor alpha production. Arthritis Rheum. (2010) 62:1213–23. 10.1002/art.27386

14 Okroj M, Heinegård D, Holmdahl R, Blom AM. Rheumatoid arthritis and the complement system. Ann Med. (2007) 39:517–30. 10.1080/07853890701477546

15 Melis L, Van Praet L, Pircher H, Venken K, Elewaut D. Senescence marker killer cell lectin-like receptor G1 (KLRG1) contributes to TNF-α production by interaction with its soluble E-cadherin ligand in chronically inflamed joints. Ann Rheum Dis. 2014;73(6):1223–1231. doi:10.1136/annrheumdis-2013-203881

16 Greenhill CJ, Jones GW, Nowell MA, et al. Interleukin-10 regulates the inflammasome-driven augmentation of inflammatory arthritis and joint destruction. Arthritis Res Ther. 2014;16(4):419. Published 2014 Aug 30. doi:10.1186/s13075-014-0419-y

